# DS4DH at MEDIQA-Chat 2023: Leveraging SVM and GPT-3 Prompt Engineering for Medical Dialogue Classification and Summarization

**DOI:** 10.1101/2023.06.08.23291121

**Authors:** Boya Zhang, Rahul Mishra, Douglas Teodoro

**Affiliations:** University of Geneva

## Abstract

This paper presents the results of the Data Science for Digital Health (DS4DH) group in the MEDIQA-Chat Tasks at ACL-ClinicalNLP 2023. Our study combines the power of a classical machine learning method, Support Vector Machine, for classifying medical dialogues, along with the implementation of oneshot prompts using GPT-3.5. We employ dialogues and summaries from the same category as prompts to generate summaries for novel dialogues. Our findings exceed the average benchmark score, offering a robust reference for assessing performance in this field.

## 1 Introduction

The unprecedented size of textual data in electronic health records has led to the information overload phenomenon (Stead and Lin, 2009), which interferes with healthcare workers’ information processing capabilities, diminishes their productivity, and prevents them from acquiring timely knowledge. Records of complex patients, such as those chronically ill, are particularly difficult to organize and to present concisely (Christensen and Grimsmo, 2008), requiring physicians to read many clinical notes during a regular medical visit, which is often unfeasible. Studies have shown that information overload can increase task demand and mental effort, which potentially impairs healthcare worker’s understanding of patients’ medical conditions and hinders optimal medical decisions, leading sometimes to fatal consequences (McDonald, 1976; McDonald et al., 2014; Karsh et al., 2006).

To tackle information overload phenomena, clinical text summarization methods have been proposed to support healthcare workers’ textual data workflow interaction (Karsh et al., 2006; Moen et al., 2016; Pivovarov and Elhadad, 2015). Clinical text summarization generates concise representations of documents using NLP methods (Manuel and Moreno, 2014). By doing so, it helps healthcare workers focus on the relevant information, which enhances medical decision-making and thus healthcare quality. Indeed, usability studies conducted with physicians for EHR summarization indicated the effectiveness of reading automatically generated summaries as compared to raw records (Wang et al., 2021).

To support efficient doctor decision-making, in this paper we investigate a novel approach that combines a traditional machine learning method, Support Vector Machines (SVM) (Cortes and Vapnik, 1995), with a cutting-edge language model, GPT-3.5 (Brown et al., 2020b), to effectively extract valuable information for the creation of doctor-patient dialogue summaries. We implemented a SVM model for short medical dialogue classification, exploring its potential on a new task to distinguish between different categories of doctor-patient encounters. Advanced generative language models have shown remarkable capabilities in text generation and reasoning. We incorporated GPT-3.5 with one-shot prompts, using dialogues and summaries from the same category as prompts to generate summaries for new dialogues. ^1^

## 2 Related Work

We discuss two key aspects of the current state of the art: (1) text classification, particularly in medical dialogue classification, and (2) summarization, with a special focus on abstractive summarization.

### Text Classification

Text classification is a well-studied problem in natural language processing, with various algorithms and techniques proposed for different domains. Traditional machine learning methods, such as Naive Bayes (John and Langley, 1995), Decision Trees (Breiman, 1984), k-Nearest Neighbors (k-NN) (Altman, 1992; Teodoro et al., 2010) and SVM (Cortes and Vapnik, 1995), have been extensively used for text classification tasks (Hartmann et al., 2019). In the medical domain, these techniques have been employed to categorize clinical notes, medical dialogues, and other types of health-related text (Obeid et al., 2019).

Deep learning approaches like Convolutional Neural Networks (CNN) (Lecun et al., 1998; Teodoro et al., 2020), Recurrent Neural Networks (RNN) (Rumelhart et al., 1986), Long Short-Term Memory Networks (LSTM) (Hochreiter and Schmidhuber, 1997), and Transformer-based architectures (Vaswani et al., 2017), including pretrained language models such as BERT (Devlin et al., 2018), RoBERTa (Liu et al., 2019), and XLNet (Yang et al., 2019), have demonstrated state-of-the-art efficacy in a diverse range of domains (Knafou et al., 2023). Leveraging the hierarchical structure of documents, graph neural networks (GNNs) have also been effectively proposed to assign categories to biomedical documents (Ferdowsi et al., 2023, 2022, 2021). Compared to deep learning models, SVM requires lower computational resources and training time and is a more efficient choice for certain applications (Sakr et al., 2016).

### Abstractive Summarization

Automatic text summarization includes extractive and abstractive summarization. Extractive summarization identifies and selects important phrases or sentences from the original text. Abstractive summarization generates summaries by creating novel sentences that capture the core information (Gupta and Gupta, 2019; Widyassari et al., 2022).

Abstractive summarization helps in generating concise representations of clinical notes, medical dialogues, and scientific articles (Joshi et al., 2020b; Cai et al., 2022). Sequence-to-sequence (seq2seq) models utilizing RNNs (Nallapati et al., 2016; Kouris et al., 2021) and Transformer architectures (Su et al., 2020; Wang et al., 2020; Laskar et al., 2022) are utilized in the abstractive summarization. The development of pre-trained language models, such as Bidirectional Encoder Representations from Transformers (BERT) (Devlin et al., 2019), Generative Pre-trained Transformer (GPT) (Brown et al., 2020a), and Text-to-Text Transfer Transformer (T5) (Raffel et al., 2020), has further advanced the state-of-the-art of this field (Ramina et al., 2020; Ma et al., 2022; Koh et al., 2022). Recent studies have explored the use of fine-tuned versions of GPT-based models for medical text summarization, showing promising results (Chintagunta et al., 2021). Our work extends this line of research by employing GPT-3.5 with one-shot prompts for medical dialogue summarization, aiming to enhance performance and practicality.

### Medical Dialogue Summarization

More recently, the summarization of medical dialogues has started to gain momentum. (Molenaar et al., 2020) use a knowledge-intensive approach, combining ontologies, guidelines and knowledge graphs to create a dialogue summarization system. The extracted triples are used to create a subjective-objective-assessment-plan (SOAP)-like report. The model achieves relatively high precision but low recall for relevant summary items. (Krishna et al., 2021) attempted the generation of complete SOAP notes from doctor-patient conversations by first extracting and clustering noteworthy utterances and then leveraging LSTM and transformer models to generate a single sentence summary from each cluster. (Joshi et al., 2020a) showed that the quality of generated summaries can be improved by encouraging copying in the pointer-generator network. Lastly, (Zhang et al., 2021) describe an abstractive approach based on BART, in which a two-stage summary model is created. The resulting models greatly surpass the performance of an average human annotator and the quality of previously published work for the task.

## 3 Methods

We address Task A of MEDIQA-Chat 2023 (Ben Abacha et al., 2023a), which focuses on Dialogue2Note Summarization in short dialogue classification and summarization. The objective of Task A is to accurately predict the summarization and section header (as shown in Table 1) for the given test set instances. The predictions are made based on the information available in the dialogue, with the token counts of the training set displayed in Figure 1.

**Table 1:** Section headers and their descriptions in medical documents.

| Label | Description |
| --- | --- |
| GENHX | General History |
| LABS | Laboratory Results |
| ROS | Review of Systems |
| FAM/SOCHX | Family and Social History |
| PASTMEDICALHX | Past Medical History |
| CC | Chief Complaint |
| ALLERGY | Allergies |
| MEDICATIONS | Medications |
| EXAM | Examination |
| PASTSURGICAL | Past Surgical History |
| ASSESSMENT | Assessment |
| IMAGING | Imaging Results |
| DIAGNOSIS | Diagnosis |
| EDCOURSE | Emergency Department Course |
| DISPOSITION | Disposition |
| IMMUNIZATIONS | Immunizations |
| GYNHX | Gynecologic History |
| PROCEDURES | Procedures |
| OTHER_HISTORY | Other History |
| PLAN | Plan |

**Figure 1:**
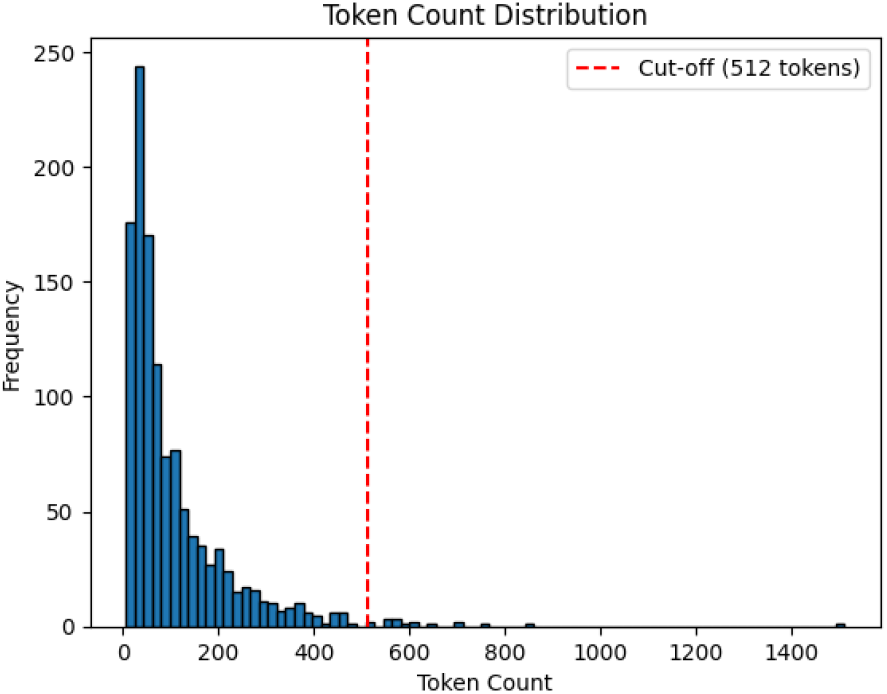
Token Count Distribution in the Dialogues.

### 3.1 Dataset

The MTS-Dialog dataset (Ben Abacha et al., 2023b) is a comprehensive and diverse collection of medical dialogues from doctor-patient encounters. We were provided with a dataset comprising 1201 training instances, 100 validation instances, and 200 test instances in the competition. Each instance in the dataset included an identifier, section header, dialogue, and summary.

### 3.2 Short Dialogue Classification

We utilized an SVM text classifier (Cortes and Vapnik, 1995) with scikit-learn (Pedregosa et al., 2011). We used CountVectorizer to transform the text into a token count matrix, considering a maximum document frequency of 0.5, a minimum document frequency of 5, and both unigrams and bigrams. Then, the token count matrix was converted into a term frequency-inverse document frequency (TF-IDF) (Salton and Buckley, 1988) representation. We employed a Stochastic Gradient Descent (SGD) (Robbins and Monro, 1951) optimization algorithm, with hinge loss, L2 penalty, and an alpha value of 1e-5. Finally, we calibrated the classifier using the Calibrated Classifier CV wrapper (Niculescu-Mizil and Caruana, 2005), enabling the provision of probability estimates.

### 3.3 Short Dialogue Summarization

#### Run 1

For the first run, we employed OpenAI’s GPT-3.5 model “gpt-3.5-turbo” ^2^ of 175 billion parameters to generate summaries based on the classified dialogues. We selected a random training instance with the same predicted section header as the instance in the test set. We then constructed three messages as input for the GPT-3.5 model.

- A user message with the content “summarize” followed by the dialogue from the selected training row.
- An assistant message containing the section text of the selected training row.
- A user message with the content “summarize” followed by the dialogue from the current test row.

The implementation was based on the OpenAI Chat API^3^ and supplied the constructed messages as input. The API returned a generated summary as part of its response.

#### Run 2

For the second run, we fine-tuned the GPT-3 curie ^4^ model (345 million parameters) on the training set. For each test instance, we extracted the dialogue text as the prompt. We used OpenAI Chat API with the fine-tuned Curie model. The output length was determined by adjusting the summary length based on the input text. We generated one completion for each input prompt with the upper limit for token length as 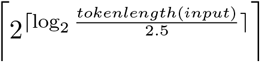. In our training dataset, the average number of tokens in the dialogue is 2.5 times greater than in the summary. We transform the upper limit to the nearest higher power of 2 by applying the base-2 logarithm.

In conclusion, both runs involved a two-stage pipeline that integrated dialogue classification and dialogue summarization, as depicted in Figure 2.

**Figure 2:**
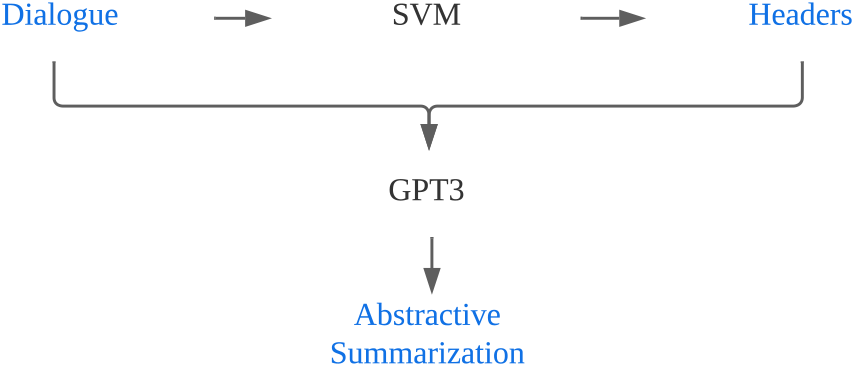
Two-Stage Pipeline for Dialogue Classification and Summarization

## 4 Experimental Results

In the following, we present the official results of our experiments on the MEDIQA-Chat 2023 Task A.

### 4.1 Short Dialogue Classification

Table 2 shows the results of our dialogue classification pipeline. Our model achieved an accuracy of 0.70. Although this result is below the best participant’s accuracy of 0.78, it surpasses the average participant’s accuracy of 0.56.

**Table 2:** Official results of MEDIQA-Chat 2023: DS4DH runs for the MEDIQA-Chat Dialogue2Note Summarization task (TaskA Header Classification).

| Run # | Accuracy |
| --- | --- |
| 1/2 | 0.70 |
| Best Participants | 0.78 |
| Average Participants | 0.56 |

### 4.2 Short Dialogue Summarization

In dialogue summarization, the perfomance of our model was evaluated using the ROUGE-1 (Lin, 2004), BERTScore F1 (Zhang and Ng, 2019), and BLEURT metrics (Sellam et al., 2020). Each evaluation metric captured different aspects of summarization quality. ROUGE-1 measures the overlap of unigrams between the generated summary and the reference summary, focusing on content similarity. BERTScore F1 evaluates the contextual embeddings of the generated and reference summaries, capturing both content and semantic similarity. BLEURT measures the summary quality by comparing the generated summary to the reference summary using a pre-trained language model, aiming to capture more complex semantic relationships. The aggregate score is calculated as the average of these three metrics.

Table 3 compares our two runs with the best and average participants’ scores across the ROUGE-1, BERTScore F1, BLEURT, and aggregate score metrics. Results show that the strategy adopted in Run 1 yields better performance compared to Run 2 (ROUGE-1: 0.3080, BERTScore F1: 0.6644, and BLEURT: 0.5206), resulting in an aggregate score of 0.4977, which also outperforms the average performance of the task participants by 2.4 percentage points. This indicates that the model provided relatively good alignment with the reference summary in terms of content, semantics, and complex relationships. Run 2 scored lower, with ROUGE-1 at 0.2937, BERTScore F1 at 0.6179, BLEURT at 0.3887, and an aggregate score of 0.4334. Nevertheless, our best model is outperformed by the top ranked run by 8 percentage points, similarly to the classification results, in which our models are also outperformed by 8 percentage points.

**Table 3:** Official results of MEDIQA-Chat 2023: DS4DH runs for the MEDIQA-Chat Dialogue2Note Summarization task (TaskA Dialogue Summarization).

| Run # | ROUGE-1 | BERTScore F1 | BLEURT | Aggregate Score |
| --- | --- | --- | --- | --- |
| 1 | **0.3080** | **0.6644** | **0.5206** | **0.4977** |
| 2 | 0.2937 | 0.6179 | 0.3887 | 0.4334 |
| Best Participants | 0.4466 | 0.7307 | 0.5593 | 0.5789 |
| Average Participants | 0.3114 | 0.6460 | 0.4630 | 0.4734 |

## 5 Discussion

### 5.1 Short Dialogue Classification

We analysed the performance of text classification model using the validation set, as ground truth labels for the test set are unavailable for post-hoc analyses. In the validation set, the model achieved a performance of 67%, which is 3% lower than the reported 70% on the test set. This discrepancy in performance can be attributed to the test set containing twice as many data points as the validation set. Despite the difference, the results imply that the model demonstrates good generalizability and avoids overfitting the training data. The relatively small performance gap between the validation and test sets suggests that the model is likely to perform well on unseen data which is a desirable trait.

Upon examining the results of the validation set as shown in the confusion matrix (Figure 3), we observe that the performance of the model was highly variable across different classes. Some classes, such as FAM/SOCHX and GENHX, showed a high degree of accurate predictions, while other classes, such as ASSESSMENT and CC, exhibited lower accuracy. This variability in performance highlights the need for further improvement and fine-tuning of the model to achieve optimal performance across all classes.

**Figure 3:**
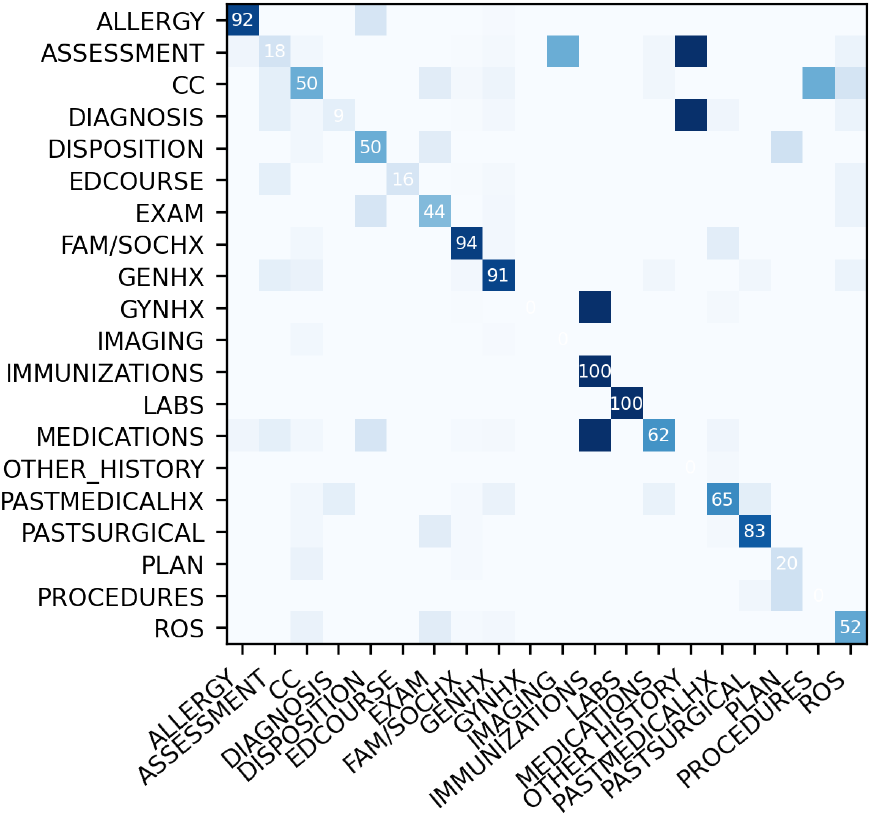
Confusion Matrix for Text Classification Model on the Validation Set

An example of the section header classifier is illustrated in Figure 4. The model displays high confidence (0.69) that the input text belongs to the “PASTMEDICALHX” (Past Medical History) class. Words such as “medical”, “diagnosis”, “conditions”, “history”, and “visit” positively contribute to the prediction. The word “medical” has the highest positive score, if omitted, the model will predict the label “PASTMEDICALHX” with a probability reduction of 0.22, leading to a confidence score of 0.47. The word “new” is negative for class “PASTMEDICALHX”. This example demonstrates the model’s ability to identify relevant keywords and distinguish between various section headers, thereby accurately classifying the input text into the appropriate category.

**Figure 4:**
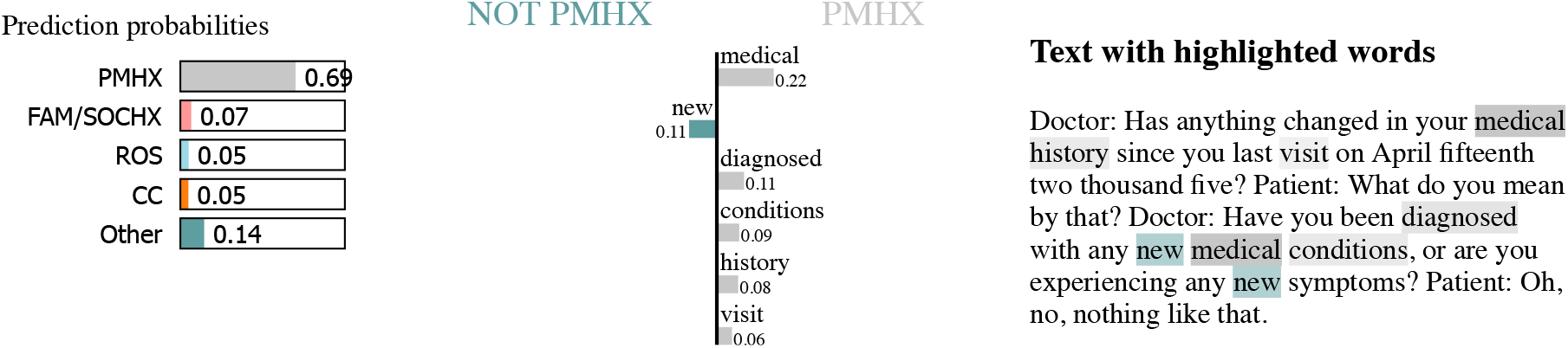
An Example for Interpreting Prediction: Header Classified as PMHX (Past Medical History)

### 5.2 Short Dialogue Summarization

#### 5.2.1 Qualitative Analyses

Table 5 displays an example in the validation set, featuring the Run 1, Run 2, and Golden summaries. These summaries are compared to evaluate their ability to effectively convey essential information.

The Run 1 summary offers a concise and clear account of the patient’s condition and history. It highlights the patient’s low back pain that started eight years ago due to a fall in an ABC store, the persistence of the pain at varying degrees, the treatments received (electrical stimulation and heat therapy), and the follow-up appointment with another doctor.

In contrast, the Run 2 summary appears less coherent, with fragmented sentences and a less organized presentation of information. It covers the fall in October 2007, pregnancy in 2008, and the worsening of back pain following another fall in 2008, but the details are not as clearly conveyed as in the Run 1 summary. Moreover, the Run 2 summary lacks clarity regarding the follow-up appointment. The Golden summary is the most comprehensive of the three, providing specific dates, treatments, and events. It outlines the patient’s history of low back pain, the treatments received, and the follow-up appointment, while also emphasizing the patient’s childbirth, which may be relevant to the case.

In conclusion, the Run 1 summary, generated by the gpt-3.5-turbo model using a single prompt and the same header class for both train and test sets, provides a concise and clear account of the patient’s situation. In contrast, the Run 2 summary, produced by the fine-tuned GPT-3 curie model using all available training data points, is less coherent and organized. This comparison highlights the potential of the gpt-3.5-turbo model to outperform the fine-tuned GPT-3 curie model, despite the latter using all available training data.

#### 5.2.2 Quantitative Analyses

Table 4 presents the results of the summarization task on the validation set, comparing the gpt-3.5-turbo ^5^ and GPT-3 curie models across various prompt strategies and evaluation metrics, including ROUGE-1, BERTScore F1, BLEURT, and an aggregate score.

**Table 4:**
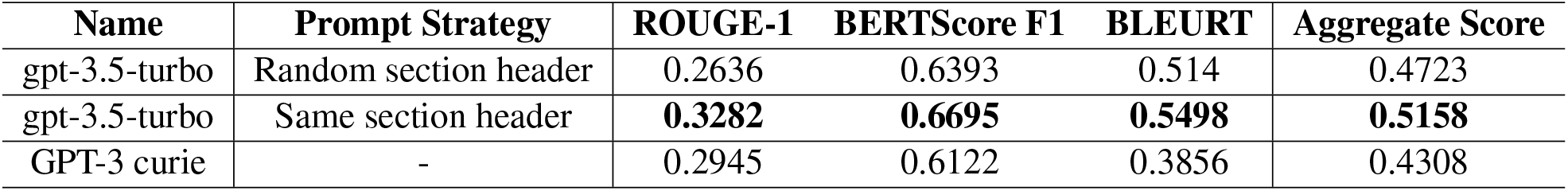
Results on the validation set for the summarization task.

**Table 5:**
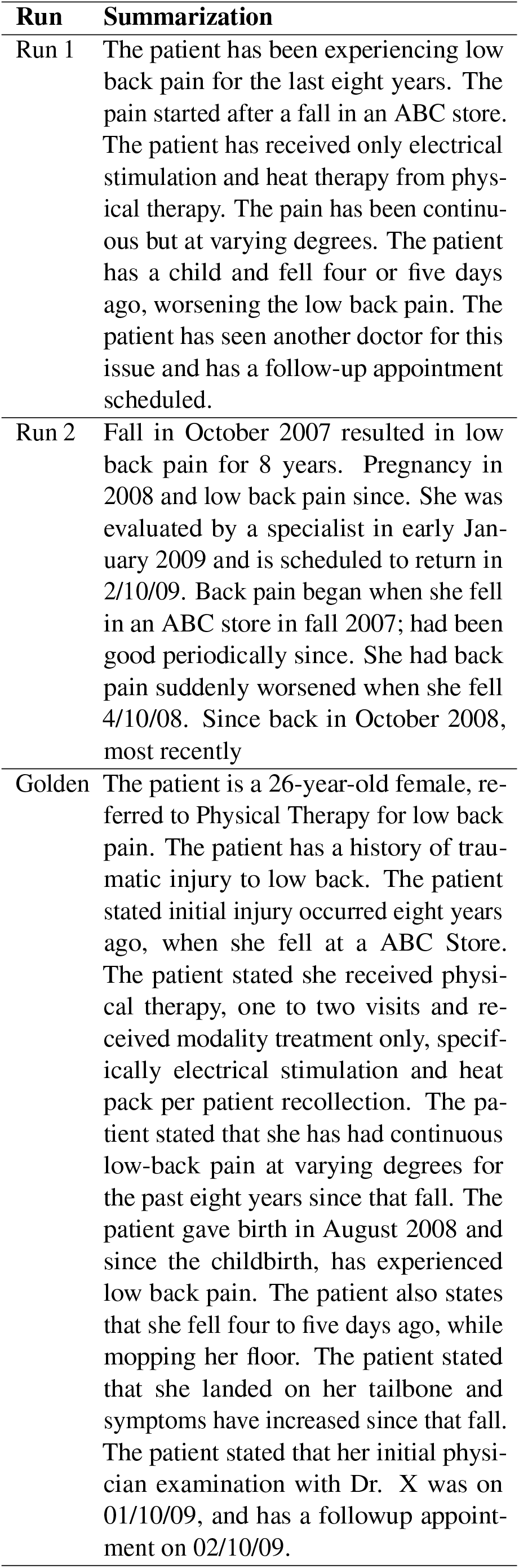
Example Summarizations: Run 1, Run 2, and Golden Summary Comparison

For the gpt-3.5-turbo model, the choice of prompt strategy significantly impacts its performance. When using a random section header as the prompt strategy, the model yields a ROUGE-1 score of 0.2636, BERTScore F1 of 0.6393, BLEURT of 0.514, and an aggregate score of 0.4723. However, by changing the prompt strategy to using the same section header, the gpt-3.5-turbo model exhibits improved performance, with a ROUGE-1 score of 0.3282, BERTScore F1 of 0.6695, BLEURT of 0.5498, and an aggregate score of 0.5158. In comparison, the GPT-3 curie model, which has been fine-tuned on the available data, achieves a ROUGE-1 score of 0.2945, BERTScore F1 of 0.6122, BLEURT of 0.3856, and an aggregate score of 0.4308. These results indicate that the gpt-3.5-turbo model, when utilizing the same section header prompt strategy, outperforms the fine-tuned GPT-3 curie model across all evaluation metrics. Furthermore, the comparison between the different prompt strategies for the gpt-3.5-turbo model highlights the importance of selecting an appropriate prompt strategy to enhance performance in the medical summarization task.

Upon comparing the oracle results from the development set with the actual results from the test set, we find that the test set results lie within the range between the upper bound (same section header) and the lower bound (random section header) of the development set. The variability within this range can be attributed to errors introduced by the classifier and helps to partially explain the gap in performance between our best model and the top-1 performance in the challenge.

### 5.3 Limitations

While our two-stage pipeline, which combines dialogue classification and dialogue summarization, has shown competitive performance compared to other participants, there are several limitations that need to be addressed for further improvement. First, both the classification and summarization tasks could benefit from enhancements in their respective models. For classification, exploring other machine learning algorithms or fine-tuning language models specifically for medical dialogue classification could potentially yield better results. Additionally, investigating the incorporation of domain-specific knowledge or leveraging external resources, like medical ontologies, might improve classification accuracy. Regarding summarization, refining the prompt strategies and experimenting with different configurations could lead to more coherent and informative summaries. This may involve exploring various prompt templates, incorporating more context from dialogues, or applying chain-of-thought reasoning to extract relevant information. Furthermore, fine-tuning the language model on a domain-specific corpus or using multi-task learning that incorporates related tasks, such as question-answering or information extraction, may contribute to better summarization performance. Finally, the evaluation metrics used in this study may not fully capture the quality of the generated summaries. It is important to acknowledge that automated evaluation metrics, like ROUGE-1, BERTScore F1, and BLEURT, might not be fully aligned with human judgments. Therefore, conducting user studies with medical professionals could provide valuable insights into the utility and accuracy of the generated summaries in real-world clinical settings.

## 6 Conclusion

Our study demonstrates the effectiveness of combining traditional machine learning techniques, such as SVM, with advanced language models, like GPT-3.5, for medical dialogue summarization. This hybrid methodology has the potential to improve documentation procedures during patient care and facilitate informed decision-making for healthcare professionals by classifying medical dialogues and generating concise summaries.

For future work, we plan to address the limitations identified in this study. For classification, we will experiment with model configurations and explore alternative machine learning algorithms. For summarization, we will refine prompt strategies, incorporate domain-specific knowledge, and investigate various fine-tuning techniques. Lastly, conducting user studies with medical professionals will provide valuable feedback to assess the utility and accuracy of our generated summaries in real-world clinical settings and further refine our approach.

## Data Availability

All data produced are available online at

https://github.com/tinaboya/MEDIQA-Chat-2023-ds4dh

## Footnotes

1 The code is available at https://github.com/tinaboya/MEDIQA-Chat-2023-ds4dh

2 https://platform.openai.com/docs/models/gpt-3-5

3 https://platform.openai.com/docs/guides/chat

4 https://platform.openai.com/docs/models/gpt-3

5 The oracle results for the GPT-3.5-turbo, in which the ground truth class is utilized for selecting the one-shot prompt, as opposed to a predicted class.

